# Quantifying the impact of introducing the HPV vaccine in 2006 on 25-29-year-old cervical cancer incidence in 2022

**DOI:** 10.1101/2025.05.20.25327999

**Authors:** Jason Semprini, Joshua Devine, Rachel Reimer

## Abstract

Nearly all cervical cancers are caused by Human Papillomavirus (HPV). In 2006, adolescent females were recommended to receive the HPV vaccine. Our study aimed to quantify the impact of introducing the HPV vaccine in 2006 on cervical cancer incidence in 2022. We analyzed the latest Surveillance, Epidemiology, and End Results data. Our design compared the change in cervical cancer incidence from 2019 to 2022 between females recommended for HPV vaccination in 2006 (age 25–29) and females who were not (age 35–54). Beyond simple pre/post comparisons, our linear regression model adjusted for age-specific incidence trends. We found that, unlike the stagnate trends in older females between 2019 to 2022, in 25-29-year-old females, cervical cancer incidence declined 2.1 cases/100,000 (CI = -2.7, -1.6): a 48% reduction from baseline trends. Although tempered by uneven adherence, after fifteen years we finally appear to be realizing quantifiable benefits from this cancer prevention vaccine.

## Introduction

Nearly all cervical cancers are caused by high-risk strains of Human Papillomavirus (HPV)^1^. In the US and globally, HPV is the most common sexually transmitted infection^2^. In the US alone, millions of people are infected, knowingly or not, with HPV each year and over 80 million Americans are infected with HPV at a given time^3^. The high transmissibility and prevalence of HPV within the US population poses a substantial challenge for cancer control systems^4^.

Although the risk of cervical cancer increases with age, cervical cancer is among the most common types of cancer in adult females under age thirty^5^. Historically, cancer control systems relied on secondary prevention methods (i.e., pap smear; HPV testing) where the goal was to identify cervical cancer at precancerous or early stages^6,7^. However, cancer control systems shifted towards primary prevention in 2006 when the HPV vaccine was introduced^8^.

Initially, only young adolescent females (age 9-13) were recommended to receive the vaccine^9,10^. In this study, we focused on the overall US population of females initially recommended to receive the HPV vaccine who have recently become young adults. It is in this cohort where we may finally observe quantifiable reductions in cervical cancer incidence because of the HPV vaccine.

## Methods

To quantify the impact of introducing the HPV vaccine in 2006 on cervical cancer incidence in 2022, we analyzed cancer incidence data from the Surveillance, Epidemiology, and End Results (SEER) program (2000-2022)^11^. The SEER-22 data included all cancer diagnoses from California, Connecticut, Georgia, Hawaii, Idaho, Illinois, Iowa, Kentucky, Louisiana, New Jersey, New Mexico, New York, Texas, and Utah as well as from the Alaska Native and Seattle registries. Incidence rates were restricted to cervical cancer only and reported as cases per 100,000 population. We excluded the years 2020-2021 to avoid potential confounding from the COVID-19 pandemic^12^. We excluded the 30-34 age group, as those females may have been influenced by the initial or ‘catch up’ HPV vaccine recommendations^9,10^.

Our design compared the change in cervical cancer incidence from 2019 to 2022 in females recommended to receive the HPV vaccine in 2006 (age 25-29 in 2022) with changes in females who were not (age 35-54 in 2022). Figure 1 visualizes our research design concept, where we leverage the age and cohort specific variation in availability of and recommendations for the HPV vaccine. If the HPV vaccine was not introduced in 2006, we’d expect cervical cancer incidence trends to remain stagnate for all age groups. Because we cannot observe this counterfactual, we compared the change in trends for the 25–29-year-old age group with trends within older age groups (35-54) and assume that in the absence of introducing the HPV vaccine, any deviations from baseline cervical cancer trends would have been similar across all age groups leading up to 2022 and beyond.

**Figure 1:**
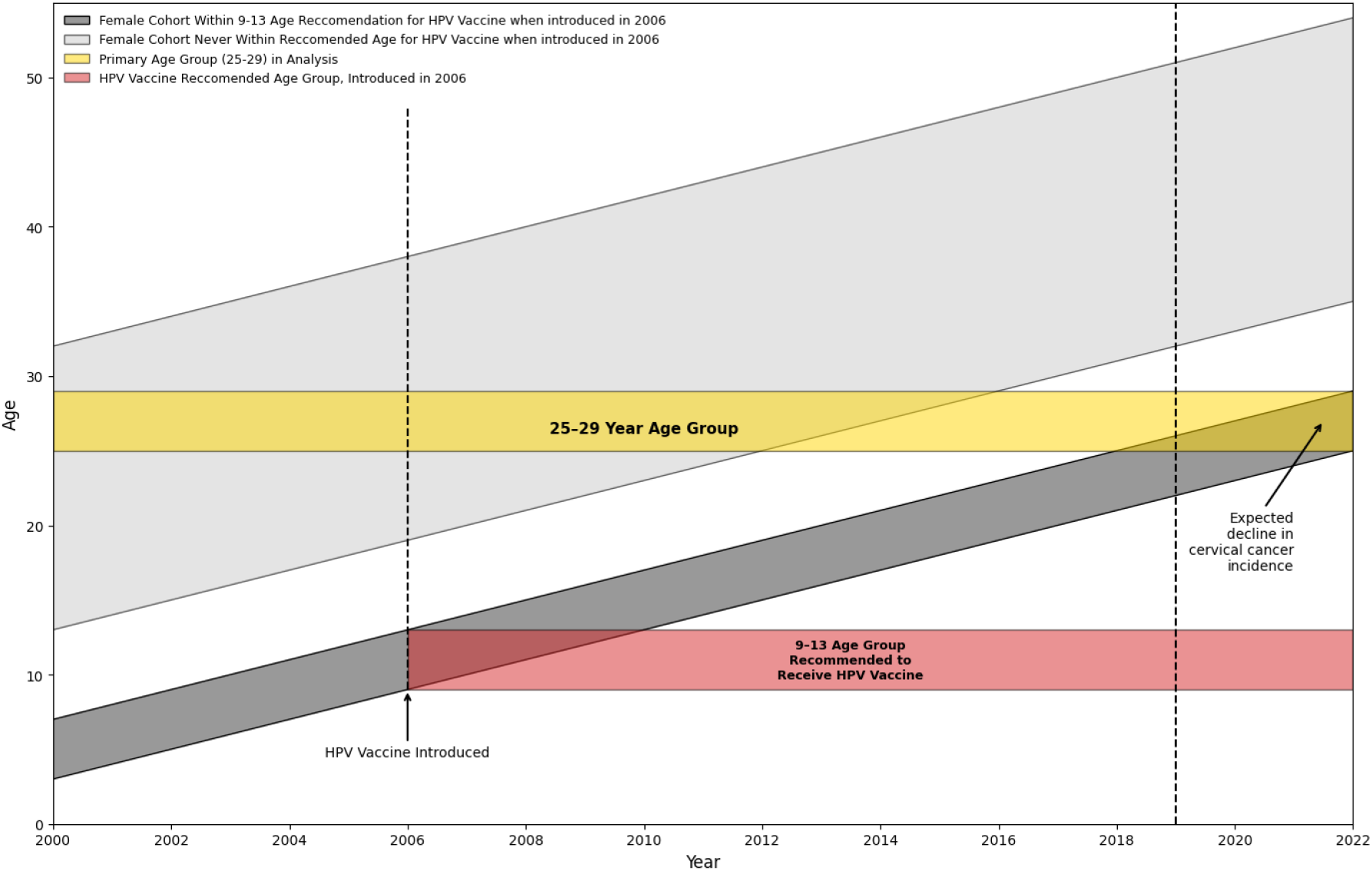
Research Design Conceptual Model. Figure 1 visualizes a conceptual model motivating our research design to quantify an expected decline in cervical cancer incidence from 2019 to 2022 among a specific cohort of 25-29-year-old females. This figure illustrates how females aged 9–13 in 2006 (gray band) fully aged into the 25–29 group (gold band) in 2022. It is in this 25-29 age group when cervical cancer incidence typically begins to rise. In 2006, the HPV vaccine was introduced and recommended for females aged 9-13 (red band). Older age groups (light gray band) were never recommended to receive the HPV vaccine and serve as comparison group. In this older comparison group, we do not expect to observe a decline in cervical cancer incidence in 2022. We only expected to observe changes in incidence among age groups who were recommended to receive the HPV vaccine in 2006 (i.e., females aged 25-29 in 2022). Conversely, we would not expect to observe changes in incidence among older age groups who were outside the age recommendation range in 2006 (i.e., 35–54-year-old females in 2022). Neither would we expect to observe changes (due to the vaccine) in cervical cancer incidence among the entire 25–29-year-old age group in years prior to 2022 (i.e., 25–29-year-olds in 2017 were 14-18 years old in 2006).

Because simple pre/post comparisons might be biased if cervical cancer incidence trends differed by age-group, our linear regression models included linear time trends and age-specific linear time trends^13^. We also tested for the presence of differential incidence trends with the 2000-2019 baseline data. Observing statistically significant differences in cervical cancer trends across age groups before 2019 would suggest changes in cervical cancer incidence may have been changing differently for reasons other than the HPV vaccine. For inference, we estimated robust bootstrapped standard errors (9,999 replications). As an additional diagnostic, we calculated formal “Difference-in-Differences” pre-trend and granger anticipation causality tests^14,15^. To determine statistical significance, we set our alpha = 0.05. All analyses were performed in Stata v. 18 (Windows). See supplemental files 1-3 for the incidence rate data, Stata do-file, and regression output.

## Results

In 2019, the cervical cancer incidence rate in 25-29-year-old females was 4.4 cases/100,000 (CI = 3.8, 5.0). In 2022, the cervical cancer incidence rate in 25-29-year-old females was 2.1 cases/100,000 (CI = 1.7, 2.6). The largest change from 2019 among older age groups was observed in 35-39-year-old females, where cervical cancer incidence dropped from 14.8 cases/100,000 to 14.2 cases/100,000. Figure 2 visualizes the cervical cancer incidence trends from 2000-2022.

**Figure 2.**
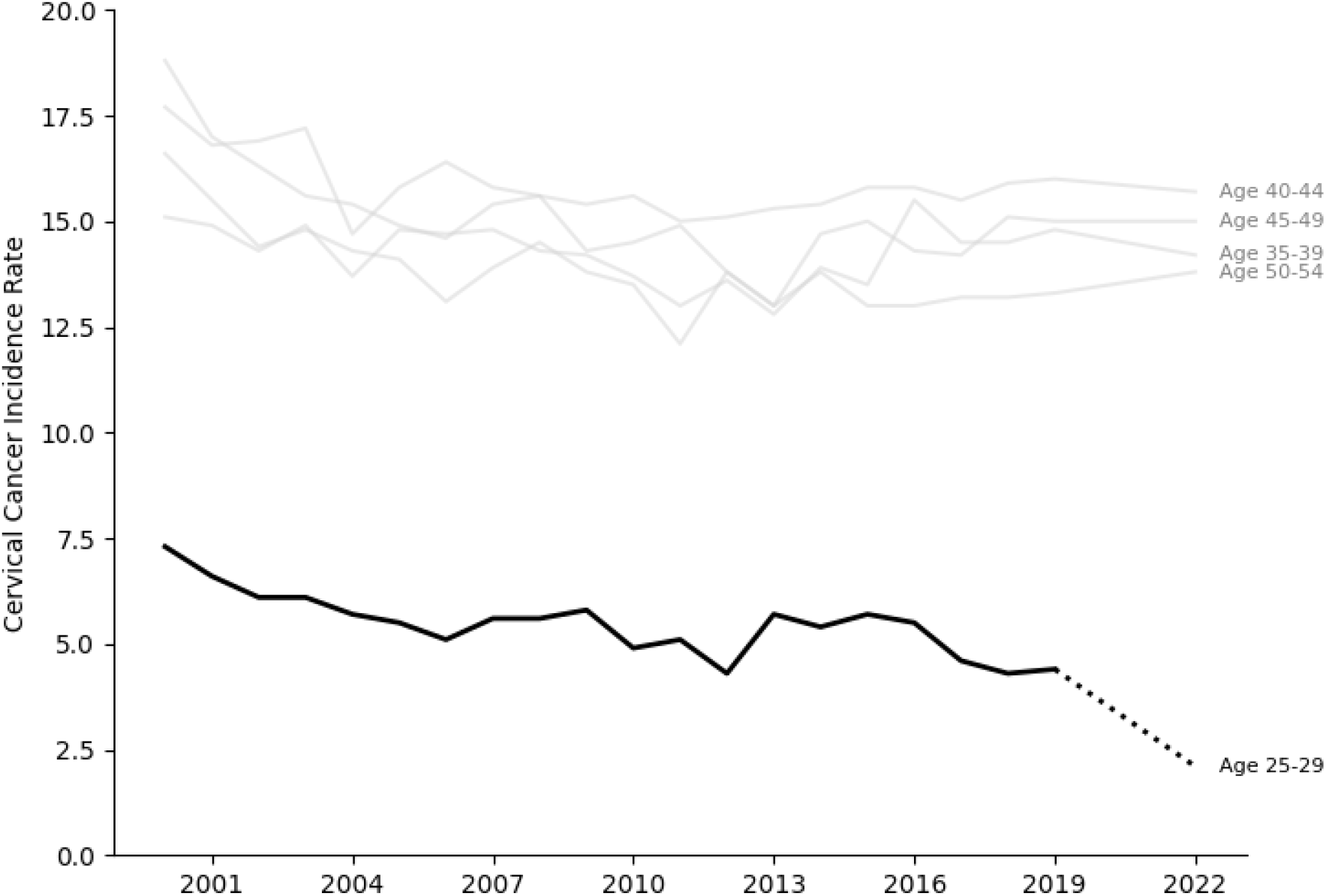
Cervical Cancer Incidence Trends. Figure 1 visualizes cervical cancer incidence rates from 2000-2019 and 2022. The dark line represents the incidence rate for females ages 25-29. The gray lines represent the incidence for females ages 35-39, 40-44, 45-49, & 50-54. The dashed line visualizes the change from 2019 to 2022 for the group recommended to receive the HPV vaccine when the HPV vaccine was introduced in 2006. All rates are per 100,000 population. Data for 2020 and 2021 were excluded to avoid confounding from the COVID-19 pandemic. Source: SEER-22.

In our main analysis we found that compared to the stagnate trends in older females, between 2019 to 2022, among 25-29-year females, cervical cancer incidence declined by 2.1 cases/100,000 (95% CI = -2.7, -1.6; p < 0.001). This decline corresponds to a 48% reduction from baseline trends. Using the 2022 population metric for registries participating in SEER (about half the cancer burden in the US), these results suggest that introducing the HPV vaccine in 2006 prevented 107 additional cervical cancers among 25-29-year-old females in 2022.

When testing for differential trends, we found that the age-specific linear trends in cervical cancer incidence were not individually or jointly different than zero. Between 2000-2019, compared to the cervical cancer incidence trend in 25-29-year-old females, the age-specific trends were as follows: 35-39 (Coef = 0.07; CI = -0.00, 0.15; p = 0.05), 40-44 (Coef = 0.03; CI = -0.05, 0.10; p = 0.44), 45-49 (Coef = -0.04; CI = -0.14, 0.07; p = 0.48), and 50-54 (Coef = -0.03; CI = -0.10, 0.04; p = 0.43). The joint test yielded a Chi2 (df =3)-test statistic of 2.3 (p=0.5131). The secondary diagnostic tests also revealed no differences in incidence rate trends prior to 2022 (Wald Pre-Trend Test p = 0.7849; Granger Test p = 0.4439). Therefore, we failed to reject the null hypothesis that the linear trends in cervical cancer incidence rates differed by age-group. Screening patterns do not explain our results (Supplemental File 4).

## Limitations

These results should be interpreted within the context of our study’s limitations. Our study did not attempt to identify any variation by individual or geographic factors, or measures of historic HPV vaccination. Future attempts to analyze subgroups should be pursued with caution, given the limited availability of HPV vaccine data linkages and sporadic incidence trends in small subpopulations. Such research, however, would overcome our overall population strategy which likely masks significant heterogeneity across the country. We also did not consider or attempt to account for the possibility of widespread HPV immunity in the 35-54 age group. Given the stagnate trends in cervical cancer incidence we observed in the study and low likelihood of completion rates in older populations, we do not consider this a major threat to our study’s design or internal validity^16^. Finally, like all non-experimental research with observational data, we cannot rule out the potential for unobserved factors correlated with age-specific cervical cancer trends unrelated to the HPV vaccine. We followed best practice testing for differential trends leading up to the year of interest, and failed to find evidence that cervical cancer incidence trends differed by age group. These empirical tests, however, can support but not confirm the validity of our research design.

## Conclusions

In 2006, adherence to the vaccine recommendation started slow and has since remained uneven^17,18^. In 2010, the proportion of females adhering to the HPV vaccine recommendation ranged from 17.6% in Idaho to 55.1% in Rhode Island^18^. Fast forward ten years and the overall proportion of females vaccinated against HPV exceeds 60%, ranging from 38-84% by state^19^. While we are pleased to observe a 48% relative drop from baseline, this is far below the potential reductions in cervical cancer we could be observing if adherence to the HPV vaccine were higher and less variable. While not yet to be realized in the US, other countries are beginning to experience the full potential of the HPV vaccine as, not just as a cervical cancer prevention tool, and elimination tool^20,21^. Whether, or when, we’ll approach such potential in the US is no guarantee.

Closing this gap represents a critical opportunity for public health. With continued efforts to improve vaccine coverage, particularly in underserved populations and low-uptake regions, we may yet accelerate the path to cervical cancer elimination. Achieving this will require not only maintaining high vaccination rates in adolescents but also addressing missed cohorts through catch-up vaccination. Importantly, the observed impact in the 25-29 age group reinforces the population-level efficacy of the vaccine, even amidst low adherence. As high resource European countries (i.e., Norway, United Kingdom) and developing countries (i.e., Rwanda) approach cervical cancer elimination thresholds, their successful HPV vaccination programs offer a blueprint for advancing progress in US^20– 24^.

As the declining incidence of cervical cancer becomes more visible, public health and cancer control systems are presented with a renewed opportunity amidst a crisis of trust in public health generally, and vaccines specifically. As a public health community, we must seize the momentum of vaccine’s success by further investing in HPV vaccine delivery across the US. Our study reaffirms the promise of the HPV vaccine as a cancer prevention tool. While possible in US, whether the next generation will realize the full potential of the HPV vaccine as a cancer elimination tool remains unclear. What must be made clear, especially to parents and guardians today, is our evidence showing that in 2022, among 25-29-year-olds residing in SEER registry areas, 109 females were diagnosed with cervical cancer and each of these 109 cancers would have been prevented by following HPV vaccine recommendations that began in 2006.

## Data Availability

See the supplemental file for the cervical cancer incidence rate data associated with this study. See the Stata do-file for analytic code to replicate the analysis and diagnostic tests.

