## Supplemental File 2 for "Quantifying the impact of introducing the HPV vaccine in 2006 on 25-29-year-old cervical cancer incidence in 2022"

clear all

**load cervical cancer incidence data (supplemental exhibit 1)*

*generate dummy variables for age groups
egen id=group(age)

*generate a binary dummy for the age 25-29 group
gen tx=0
replace tx=1 if id==1

*generate a binary "exposed" variable (interaction between age 25-29 & year == 2022)
gen expose=0
replace expose=1 if tx==1 & year==2022

***baseline coefficient pre-trend test***
regress rate c.year##ib1.id if year<2020, vce(bootstrap, reps(9999) seed(123) dots(500))

test 3.id#c.year = 4.id#c.year = 5.id#c.year = 0

***primary regression model - linear regression adjusting for linear trends and age-specific linear trends
reg rate i.expose c.year##i.id , vce(bootstrap, reps(9999) seed(123) dots(500))

*compute event history analysis
reg rate i.year##i.tx, vce(robust)

*perform diagnostic difference-in-difference analysis to formally test pre-trends and granger causality - NOTE: this regression may not yield an appropriate estimate of impact on cervical cancer incidence as this Dif-in-Dif approach does NOT allow for age-specific trends in cervical cancer incidence like our primary model*

didregress (rate) (expose), group(id) time(year) 
estat ptrends 
estat granger
estat trendplots
estat grangerplot
