## Supplemental File 3 for "Quantifying the impact of introducing the HPV vaccine in 2006 on 25-29-year-old cervical cancer incidence in 2022"

**Supplemental File 3 – Diagnostic Results**

**Supplemental File 3a – Testing for Differences in Cervical Cancer Trends (2000-2019), by age group**

| rate | Observed coefficient | Bootstrap std. err. | z | P>\|z\| | Normal-based [95% conf. | interval] |
|---|---|---|---|---|---|---|
| year | -.0978195 | .0205961 | -4.75 | 0.000 | -.1381872 | -.0574519 |
| id | | | | | | |
| 2 | -133.8192 | 72.71641 | -1.84 | 0.066 | -276.3407 | 8.702379 |
| 3 | -48.50517 | 76.70263 | -0.63 | 0.527 | -198.8396 | 101.8292 |
| 4 | 85.35117 | 106.1342 | 0.80 | 0.421 | -122.668 | 293.3703 |
| 5 | 67.00303 | 74.30646 | 0.90 | 0.367 | -78.63495 | 212.641 |
| id#c.year | | | | | | |
| 2 | .0709774 | .0362151 | 1.96 | 0.050 | -2.84e-06 | .1419577 |
| 3 | .0293233 | .0381483 | 0.77 | 0.442 | -.045446 | .1040926 |
| 4 | -.0376692 | .0527974 | -0.71 | 0.476 | -.1411502 | .0658119 |
| 5 | -.0291729 | .0369583 | -0.79 | 0.430 | -.10161 | .0432641 |
| _cons | 202.0334 | 41.38552 | 4.88 | 0.000 | 120.9192 | 283.1475 |

Supplemental File 3a estimates a linear regression model (via bootstrap):

Rate ~ c.year + i.$AGE^{id}$ + $AGE^2$#c.year + $AGE^3$#c.year + $AGE^4$#c.year+ $AGE^5$#c.year

Individually, the linear trend of cervical cancer incidence for each age group did not statistically significantly differ from the linear trend observed in the primary age group of interest (Age ID 1 = 25–29-year-old).

**Supplemental File 3b – Testing for Joint Differences in Cervical Cancer Trends (2000-2019)**

```
( 1)  3.id#c.year - 4.id#c.year = 0
( 2)  3.id#c.year - 5.id#c.year = 0
( 3)  3.id#c.year = 0

          chi2(  3) =    2.30
        Prob > chi2 =    0.5131
```

After estimating the regression model from Supplemental File 3a, we tested if each age-specific linear trend coefficient were jointly equal to zero. We fail to reject the null hypotheses that the age-specific linear trends were statistically different leading up to 2019.

**Supplemental File 3c – Primary Regression Result Table**

| rate | Observed coefficient | Bootstrap std. err. | z | P>\|z\| | Normal-based [95% conf. | interval] |
|---|---|---|---|---|---|---|
| 1.expose | -2.142256 | .2864745 | -7.48 | 0.000 | -2.703735 | -1.580776 |
| year | -.0978195 | .0209912 | -4.66 | 0.000 | -.1389615 | -.0566776 |
| id | | | | | | |
| 2 | -141.4674 | 63.04292 | -2.24 | 0.025 | -265.0293 | -17.90556 |
| 3 | -68.20963 | 71.33185 | -0.96 | 0.339 | -208.0175 | 71.59822 |
| 4 | 39.15484 | 101.6166 | 0.39 | 0.700 | -160.0101 | 238.3198 |
| 5 | 21.72271 | 78.32569 | 0.28 | 0.782 | -131.7928 | 175.2382 |
| id#c.year | | | | | | |
| 2 | .0747885 | .0313879 | 2.38 | 0.017 | .0132694 | .1363077 |
| 3 | .0391419 | .0354665 | 1.10 | 0.270 | -.030371 | .1086549 |
| 4 | -.0146497 | .0505373 | -0.29 | 0.772 | -.113701 | .0844016 |
| 5 | -.00661 | .0389581 | -0.17 | 0.865 | -.0829665 | .0697466 |
| _cons | 202.0334 | 42.18171 | 4.79 | 0.000 | 119.3587 | 284.708 |

Supplemental File 3c estimates a linear regression model:

Rate ~ EXPOSE + c.year + i.$AGE^{id}$ + i.$AGE^{id}$#c.year

This regression model estimates the association between the EXPOSE variable and cervical cancer incidence rate. EXPOSE is a binary variable, taking the value of 1 if Age = 25-29 and year = 2022 (zero else). The coefficient on the EXPOSE variable quantifies (estimates) the impact of introducing the HPV vaccine in 2006 on cervical cancer incidence in 2022 among the primary age group (25-29). This regression adjusts for global linear time trends, age-specific baselines, and age-specific trends in cervical cancer incidence.

**Supplemental File 3d – Event-History Plot**

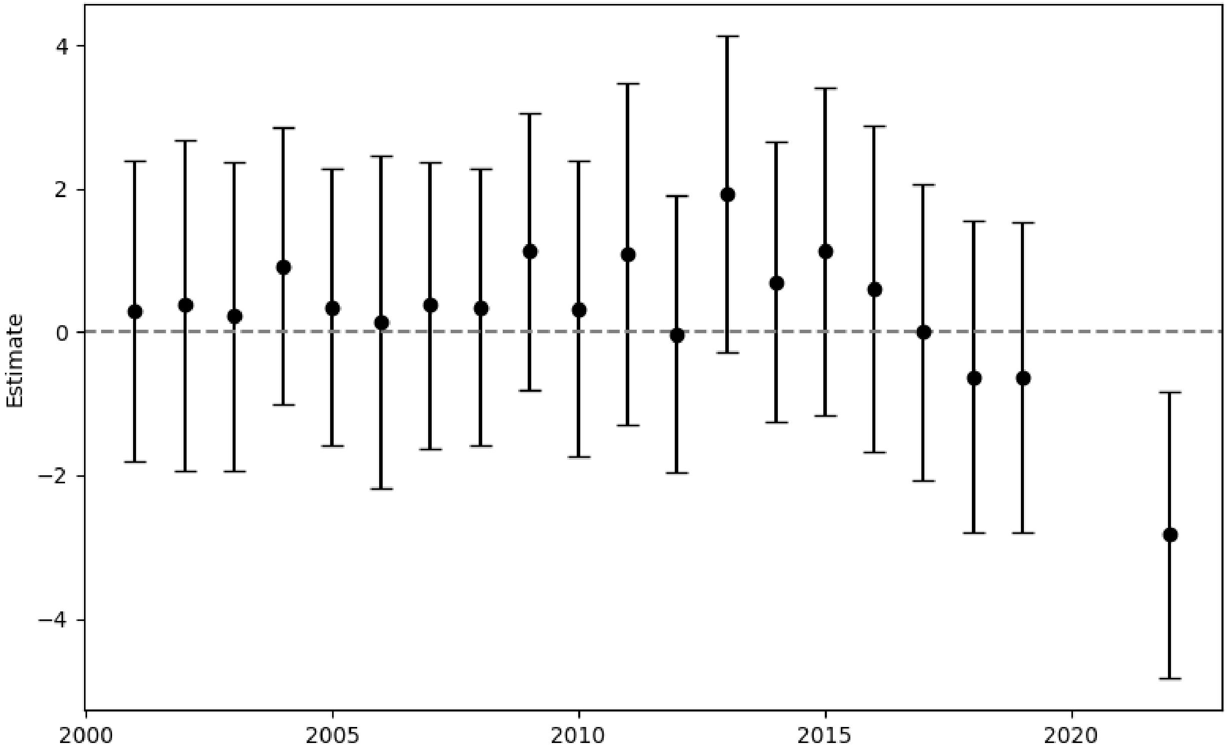


Supplemental File 3d visualizes the estimated year-by-year differences in the 25-29-year-old cervical cancer incidence rate and the average cervical cancer incidence rate among 35–54-year-olds. The error bars represent 95% confidence intervals. The event history analysis and confidence intervals were estimated via linear regression. All nineteen baseline (pre-2022) estimates were not statistically different than zero, suggesting that cervical cancer incidence trends were consistent between age groups until 2022.

**Supplemental File 3e – Formal Difference-in-Differences Diagnostics**

```
Parallel-trends test (pretreatment time period)
H0: Linear trends are parallel

 F(1, 4) =   0.09
Prob > F = 0.7849

Granger causality test
H0: No effect in anticipation of treatment

 F(3, 4) =   1.11
Prob > F = 0.4439
```

After estimating a Difference-in-Differences regression in Stata v. 18 (command didregress), supplemental 3e reports the post-estimation diagnostic tests. First, the "Paralell-trends" test, tests for differences in cervical cancer incidence trends between the primary age group (25-29) and the average trend in older groups (35-54). This result suggests that we failed to reject the null hypothesis of paralell-trends. The "Granger Causality" or anticipation test, tests if cervical cancer incidence trends began to diverge leading up to the year 2019 (instead of after) in the primary age group (25-29). This result suggests that we failed to reject the null hypothesis that there was no anticipatory difference in cervical cancer incidence trends before 2019.

**Supplemental File 3f – Average Observed and Modelled Trends in Cervical Cancer Incidence**

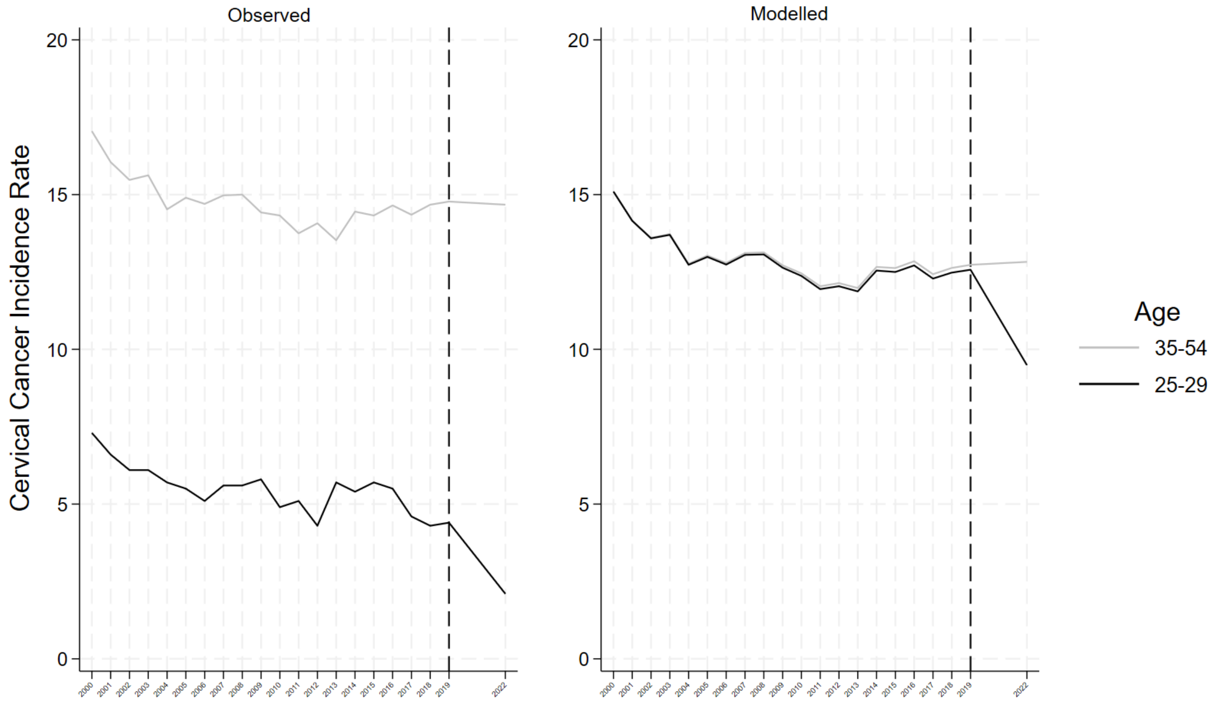


After estimating a regression model, supplemental 3f visualizes the observed mean incidence rates by age group categories (25-29 and 35-54) and a modelled rate. The left panel is straightforward, where the 25-29 observed rate is the actual rate for each year and the 35-54 observed rate is the mean rate for each older age group. The right panel is derived from a post-estimation proceedure, where after regression, the figure displays model-based predictions from an augmented regression that interacts age-group with each year. This allows us for year-specific deviations in the primary age group's outcome trend. This produces predicted values for both treatment and control groups under a model that adjusts for pre-period trends. In this context, the divergence between the 25–29 and 35–54 age groups after the dashed vertical line (marking the post-treatment period) reflects adjusted, model-implied differences in cervical cancer incidence, accounting for prior patterns in each group.
