## Supplemental File 4 for "Quantifying the impact of introducing the HPV vaccine in 2006 on 25-29-year-old cervical cancer incidence in 2022"

**Supplemental File 4- Cervical Cancer Screening (Pap Smear) Rates**

If cervical cancer screening rates varied differently over time across age groups, our design estimating the effect of introducing the HPV vaccine in 2006 on cervical cancer incidence in 2022 may be biased. Cervical cancer screening rates could change over time differently by age for a number of reasons. Perhaps introducing the HPV vaccine raised screening awareness among age groups who were NOT recommended to complete a screening. Moreover, the potential bias from differential trends in cervical cancer screening rates across age is ambiguous. First, higher screening rates could lead to higher incidence due to improved detection patterns. Conversely, higher screening rates could lead to lower incidence due to removing precancerous lesions before malignancy. To assess the extent to which this potential confounding biases our estimates, we first tested for differences in cervical cancer screening trends and then re-estimated our primary regression model adjusting for annual cervical cancer screening rates.

We accessed individual-level survey data from the Behavioral Risk Factor Surveillance System (BRFSS). Survey years included all modules with cancer screening questions (2000, 2002, 2004, 2006, 2008, 2010, 2012, 2014, 2016, 2018, 2020, 2022). Using two survey questions, we created a variable indicating if the respondent had ever received a pap smear and, if so, received that pap smear in the past year. After visualizing sampling weighted average trends for each age group, we tested if average pap smear screening trends varied by age group with a linear regression model. Then, we merged the pap smear data to the cervical cancer incidence rate data. With this merged data, we replicated our primary estimate to assess if the results changed after including continuous measure of pap smear rates as a control variable.

Here is the age-stratified figure showing declining trends in annual cervical cancer screening (pap smear).

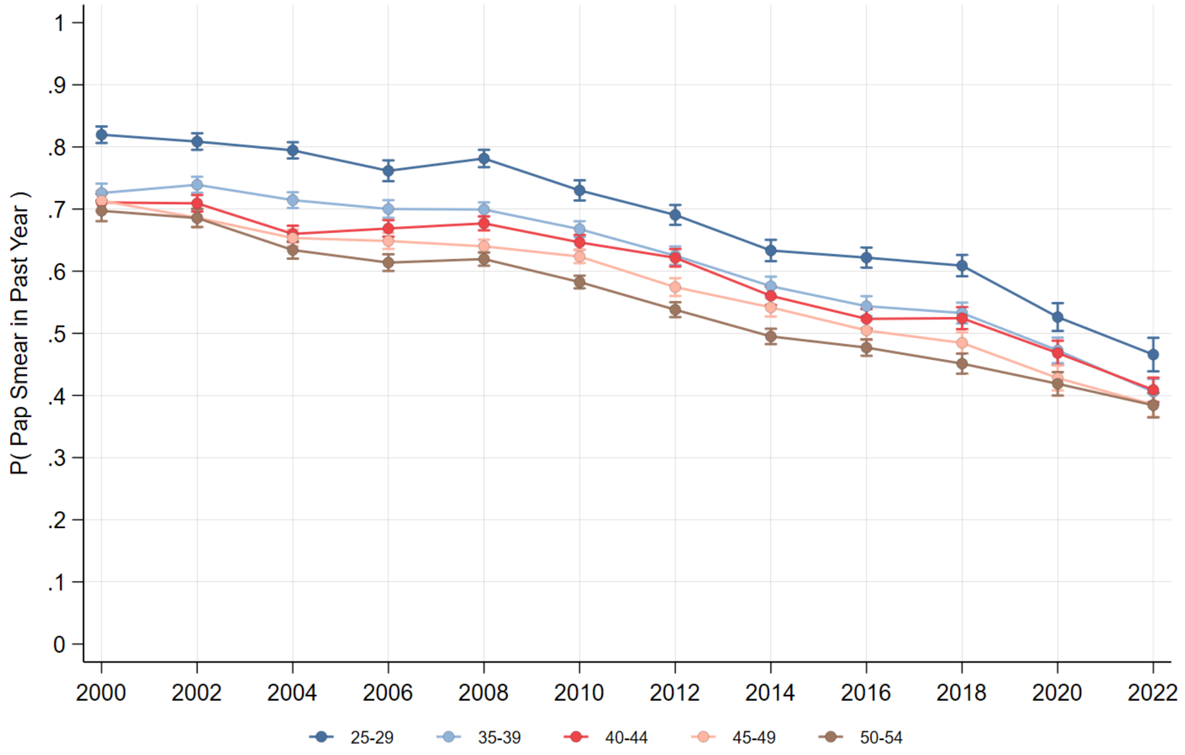

Here is the table showing the results of the differential trend tests (including all years). We fail to reject the null hypothesis that pap smear trends changed differently across age groups.

| pap_yr | Observed coefficient | Bootstrap std. err. | z | P>\|z\| | Normal-based [95% conf. | interval] |
|---|---|---|---|---|---|---|
| year | -.0155869 | .0017267 | -9.03 | 0.000 | -.0189712 | -.0122026 |
| id | | | | | | |
| 2 | -1.940115 | 5.094557 | -0.38 | 0.703 | -11.92526 | 8.045033 |
| 3 | -4.934567 | 4.710012 | -1.05 | 0.295 | -14.16602 | 4.296887 |
| 4 | -2.706577 | 4.246317 | -0.64 | 0.524 | -11.0292 | 5.616051 |
| 5 | -2.565246 | 3.659503 | -0.70 | 0.483 | -9.737741 | 4.607248 |
| id#c.year | | | | | | |
| 2 | .0009299 | .0025335 | 0.37 | 0.714 | -.0040356 | .0058954 |
| 3 | .0024097 | .0023424 | 1.03 | 0.304 | -.0021813 | .0070007 |
| 4 | .0012896 | .0021119 | 0.61 | 0.541 | -.0028497 | .0054289 |
| 5 | .0012074 | .0018199 | 0.66 | 0.507 | -.0023596 | .0047744 |
| _cons | 32.03216 | 3.471445 | 9.23 | 0.000 | 25.22825 | 38.83606 |

```
. test 2.id#c.year = 3.id#c.year = 4.id#c.year = 5.id#c.year = 0

 ( 1)  2.id#c.year - 3.id#c.year = 0
 ( 2)  2.id#c.year - 4.id#c.year = 0
 ( 3)  2.id#c.year - 5.id#c.year = 0
 ( 4)  2.id#c.year = 0

           chi2(  4) =    1.09
         Prob > chi2 =    0.8952
```

Adding the average annual pap smear screening rate to the primary regression model does not differ in any statistical or meaningful way. Note, the model with the pap smear control variable only includes even years, effectively estimating the change in cervical cancer incidence for 25-29-year-old females from 2018 to 2022. While our primary model estimated the between 2019 to 2022, cervical cancer incidence in this age group declined by 2.1 cases/100,000 (95% CI = -2.7, -1.6; $p < 0.001$). Adjusting for annual pap smear screening rates, we estimated that between 2018 to 2022 cervical cancer incidence declined 2.9 cases/100,000 (CI = -4.3, -1.5; $p < 0.001$).
